# A nonrandomized phase 2 trial of EG-Mirotin, a novel, first-in-class, subcutaneously delivered peptide drug for non-proliferative diabetic retinopathy

**DOI:** 10.1101/2021.12.30.21267814

**Authors:** Seunghoon Yoo, Dae Hyuk You, Jeongyoon Lee, H. Christian Hong, Sung Jin Lee

## Abstract

**Aim:** EG-Mirotin (active ingredient EGT022) targets non-proliferative diabetic retinopathy (NPDR), the early stage of retinopathy. EG-Mirotin reverses capillary damage before NPDR progresses to an irreversible stage. EG-Mirotin safety and efficacy were investigated in patients with type 1 or type 2 diabetes mellitus and moderate-to-severe NPDR.

**Methods:** In this open-label, single-arm, single-center, exploratory phase II study, 10 patients (20 eyes) received EG-Mirotin once a day (3 mg/1.5 ml sterile saline) for 5 days and were evaluated for ischemic index changes and safety. End-of-study was approximately 8±1 weeks (57±7 days) after the first drug administration.

**Results:** EG-Mirotin injections were well tolerated with no dose-limiting adverse events, serious adverse events, or deaths. Four treatment-emergent adverse events (TEAEs) unrelated to the investigational drug were observed in 2 out of 10 participants (20.00%) who had received the investigational drug. The overall average percent change in ischemic index at each evaluation point compared to baseline was statistically significant (Greenhouse-Geisser F=9.456, p=0.004 for the main effect of time), and a larger change was observed when the baseline ischemic index value was high (Greenhouse-Geisser F=10.946, p=0.002 for time × group interaction).

**Conclusions:** EG-Mirotin was well tolerated and reversed diabetes-induced ischemia and leakage of capillaries in the retina.

## 1. INTRODUCTION

Even in the 21^st^ century, diabetes mellitus is rapidly spreading as a global epidemic. Currently, 463 million adults (20–79 years) are affected worldwide, and this number will reach 700 million by 2045 [1]. Diabetic retinopathy (DR), a disease complication, is the leading cause of blindness in people aged 20–74 years [2]. DR is caused by diabetes-induced injury of the retinal microvasculature. It is classified as non-proliferative DR (NPDR) or proliferative DR (PDR) [3]. Furthermore, diabetic macular edema (DME) can cause severe visual impairment [4].

Diabetes damages endothelial cells [5], which stimulates cell proliferation and motility. Microvessels are formed by vascular endothelial cells and pericytes that support and surround them [6,7]. Pericytes contribute to the stabilization of capillary beds, and their correct interaction with the extracellular matrix is essential for microvascular development [8]. If diabetes-induced changes worsen, pericyte loss and basement membrane thickening will eventually lead to loss of microvessels, i.e., NPDR [9]. These vascular abnormalities may lead to inadequate blood flow in retinal areas. To adjust for hypoperfusion, angiogenesis occurs causing undesired neovascularization, retinal detachment, and eventually loss of sight. In patients with diabetes mellitus for more than 20 years, an estimated 86% and 77% of patients with type 1 and type 2 diabetes, respectively, experience loss of vision [10]. Moreover, patients with DR contribute disproportionately to healthcare and societal costs caused by diabetes mellitus.

DR prevention consists primarily of the optimal control of blood glucose levels [11], as well as the treatment of other risk factors, especially hypertension and hyperlipidemia. However, no appropriate therapy for early-stage retinopathy (NPDR) and only limited treatment options for late-stage retinopathy (PDR) exist. Once apparent, PDR can be treated by laser coagulation and intraocular injection of anti-angiogenic drugs, such as bevacizumab, ranibizumab, and pegaptanib [12]. Current DR treatment involves injecting anti-vascular endothelial growth factor (anti-VEGF) antibodies to inhibit angiogenesis in the ischemic retina and prevent vascular leakage caused by pericyte loss. However, this treatment cannot restore the retina because capillaries are irreversibly damaged. To improve outcomes, new treatment paradigms are required. It is necessary to delay vision loss as much as possible by preventing damage to capillaries or restoring them in the NPDR stage.

EyeGene, Inc. developed the new drug EG-Mirotin that contains the active ingredient EGT022. EG-Mirotin targets NPDR before capillary damage progresses to an irreversible stage and offers a completely new treatment approach. Most patients with DR experience anxiety when they hear that the drug is injected into their eyes. However, the main advantage of EG-Mirotin is that it reduces patient anxiety because it can be injected subcutaneously with a small-gauge (31–29) needle.

EGT022, a recombinant polypeptide derived from human a disintegrin and metalloproteinase (ADAM)15, contains 58 amino acids and weighs 6.1 kDa. After subcutaneous injection of EGT022, it binds to integrins on the surface of thrombocytes, stimulating their production of growth factors including angiopoietin 1. In DR, capillaries are prone to rupture and subsequent retinal hemorrhages and fluid leakages, resulting in retinal and macular edemas [13]. The primary activity of thrombocyte-released growth factors, especially angiopoietin 1, following EGT022 administration appears to be the recruitment and development of pericytes, specialized connective tissue cells, which together with endothelial cells constitute capillaries [14].

EG-Mirotin successfully completed phase 1 clinical trials demonstrating excellent safety. The current trial aimed to evaluate the efficacy and safety of subcutaneous EG-Mirotin injection in adult NPDR patients with diabetic edema.

## 2. Materials and methods

### 2.1. Study drug

EG-Mirotin was provided by EyeGene, Inc. (Seoul, Korea) as a freeze-dried powder. EGT022 was cloned using the N-terminal primer 5’-CTCGAGAAAGAACCTGCCAGCTGA-3’ and the C-terminal primer 5’-GCGGCCGCTCAGCCATCCCCTAGGC-3’ from the human fetal liver cDNA library. We used the previously described *Pichia pastoris* expression system [15].

### 2.2. Trial design

This was an open-label, single-arm, single-center, exploratory phase II study. The protocol was approved by the Ministry of Food and Drug Administration of Korea (approval no. 32764) and by the investigational review board of Soonchunhyang University Hospital. Patients gave written consent after being informed about the characteristics of the clinical trial and investigational drug.

### 2.3. Study population

We enrolled 10 patients (8 men, 2 women) with moderate-to-severe NPDR.

**Table 1.** Patient demographics and main clinical findings

|  |  |
| --- | --- |
| Patients/Eyes | 10/20 |
| Age | 61.50± 0.17 |
| Sex(Male/Female) | 8/2 |
| VA1 | 0.89±0.17 |
| CMT1 | 288.90±21.05 |
| RV1 | 0.23±0.02 |
| Total Ischemic Retina | 793.00±20.03 |
| Non-Perfused Retina | 29.78±36.33 |
| Ischemic Index | 2.12±4.78 |
| Total Leakage Retina | 796.00±21.62 |
| Capillary Leakage | 108.65±90.26 |
| Leakage Index | 13.595±11.21 |

VA1: Visual acuity; CMT 1: Continuing medication and treatment; RV 1: Residual volume

#### 2.3.1. Inclusion criteria

The inclusion criteria were: (i) age >19 years; (ii) presence of type 1 or type 2 diabetes according to the following criteria: patients fulfill the American Diabetes Association or World Health Organization criteria, patients regularly take insulin or receive parenteral treatment for diabetes, or patients regularly take oral hypoglycemic agents for diabetes treatment; (iii) moderate-to-severe NPDR (e.g., Early Treatment Diabetic Retinopathy Study (ETDRS) severity 43A-53) for the study eye; (iv) the investigator considers that photocoagulation and anti-VEGF treatment can be safely postponed until visit (V)9 (8 weeks); and (v) media clarity and pupillary dilation is maintained as far as fundus imaging allows, allowing adequate evaluation of reference fundus imaging results, and subject for the case of ophthalmology and other examinations.

#### 2.3.2. Exclusion criteria

Patients were excluded if any of the following criteria applied to the study eye: (i) glaucoma and glaucoma-related disorders, retinal detachment, macular degeneration; (ii) cataract and vitreous hemorrhage; (iii) eye diseases other than DR affecting the evaluation (e.g., retinal vein occlusion, uveitis, other inflammatory ophthalmic diseases, eye tumors); (iv) new blood vessels in the disc, retina, iris, or angle; (v) fluorescein angiography showing macular ischemia, ≥grade 2; (vi) panretinal photocoagulation (PRP) or scheduled PRP prior to V9; (vii) triamcinolone acetonide intravitreal and peribulbar steroid injections within 6 months of clinical trial drug administration, vitrectomy, or intravitreal injection of anti-VEGF drugs; (viii) capsulotomy (e.g., yttrium-aluminum-garnet capsulotomy) within 2 months prior to enrollment; (ix) major ophthalmic surgery within 3 months (e.g., cataract, glaucoma) or scheduled surgery prior to V9; (x) acquired aphakia as congenital, surgical, and traumatic; and (xi) untreatable intra- or periocular infection.

Systemic exclusion criteria were: (i) difficult glycemic control, so patients received or planned to receive intensive insulin therapy; (ii) uncontrolled hypertension; (iii) clinically significant arrhythmias; (iv) history of myocardial infarction in the last 3 months; (v) severe kidney failure including dialysis; (vi) severe liver failure; (vii) history of malignancy within 3 years, except for thyroid cancer (papillary, follicular, or medullary types corresponding to stage I or II), basal cell or squamous cell carcinoma of the skin, cervical intraepithelial neoplasia (CIN) and carcinoma in situ (CIS) of the cervix, and intraepithelial carcinomas in other organs; (viii) immunodeficiency; (ix) autoimmune diseases (e.g., multiple sclerosis, myasthenia gravis, acute disseminated encephalomyelitis); (x) history of hypersensitivity reaction to EG-Mirotin components; (xi) history of fluorescein hypersensitivity; (xii) patients received contraindicated drugs or treatments before participating in this trial and have not passed the prescribed period (data not shown); (xiii) patients took contraindicated drugs (data not shown); (xvi) pregnant or breastfeeding women; (xv) women (age<50 years) who were not permanently infertile and a pregnancy was not excluded by a pregnancy test; (xvi) refusal to accept an effective contraceptive method during the clinical trial period; (xvii) current participation in another clinical trial or patients who have not passed 30 days from the date of signature of the consent form after participating in the previous clinical trial, and (xix) diseases not specified above.

### 2.4. Drug administration

Enrolled patients received EG-Mirotin once per day (3 mg in 1.5 ml of sterile saline) for 5 days (five times in total) with an additional follow-up period of approximately 7 weeks (8 weeks in total: screening [V1], treatment period) [V2–V6], follow-up period [V7–V9]). End-of-study was approximately 8±1 weeks (57±7 days) after the first dose.

### 2.5. Evaluation of the ischemic index and leakage index

Ischemic index and leakage index values were measured using ultra-wide-field fluorescein angiography (UWFA) (Fig. 1) with an Optos California (Optos PLC, Dunfermline, UK) for ultra-wide angiography and ultra-wide fluorescence angiography, respectively. To determine the indexes, total retinal area, ischemic area, and leakage area were measured using the Optos Advance software (Optos PLC). On two UWFA images obtained at approximately 40 s and 8 min after contrast medium injection, one objective evaluator marked the maximal identifiable retina with a continuous line to calculate total retinal areas (in mm^2^). Summed retinal areas marked by the assessor as non-perfused regions in the first image and leaking capillaries in the second image were defined as ischemic and leakage areas, respectively. The ischemic and leakage indexes (in %) were defined as the ratios of ischemic and leakage areas, respectively, divided by the total retinal area. Total retinal areas may differ within an individual, but the total area values remained unchanged throughout the study.

**Fig. 1.**
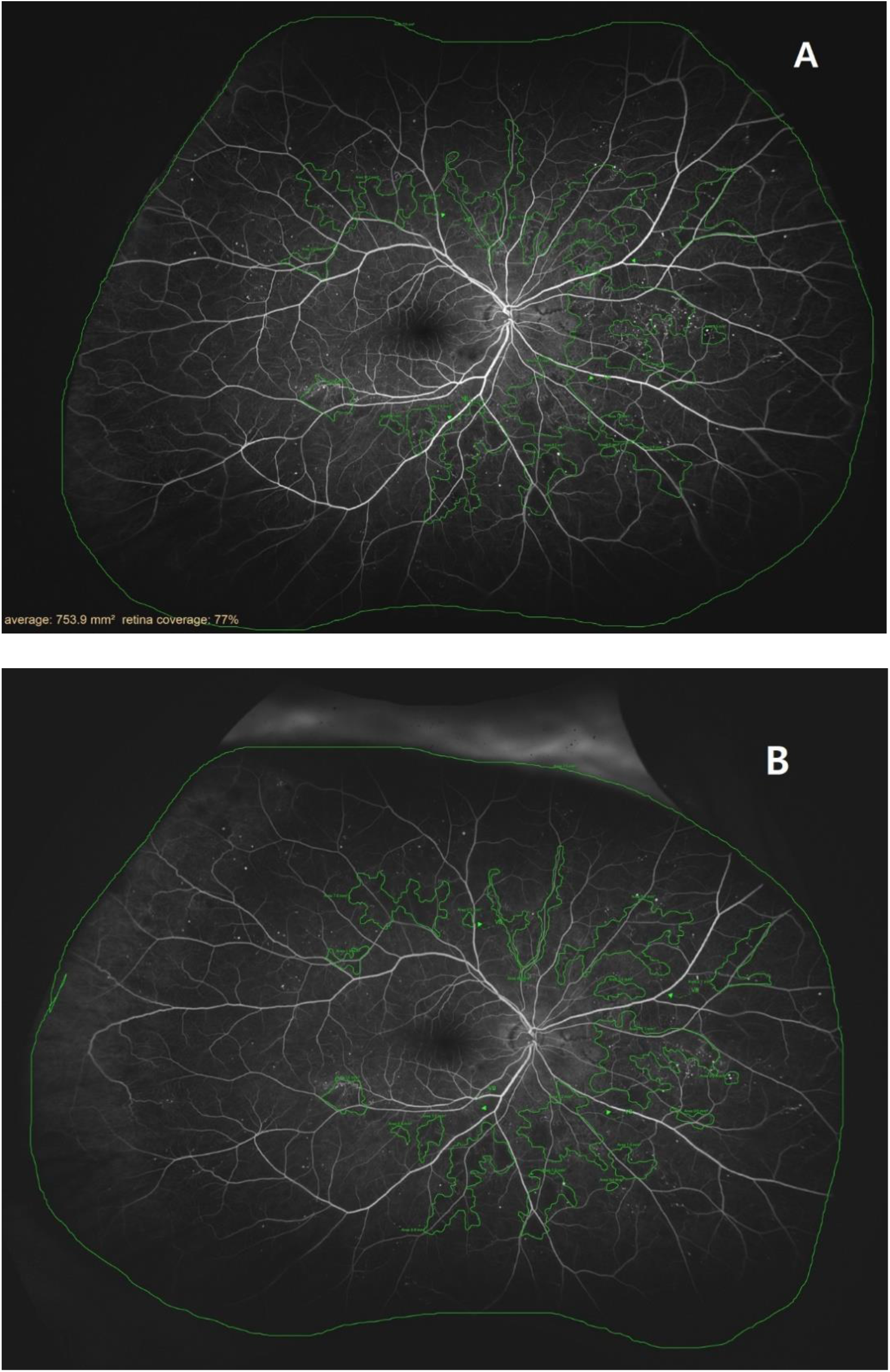
Example tracing images of retinas examined with ultra-wide-field fluorescein angiography. (A) A retina 2 weeks before the treatment (V1). (B) A retina 8±1 weeks after the first treatment (V9).

**Fig. 2.**
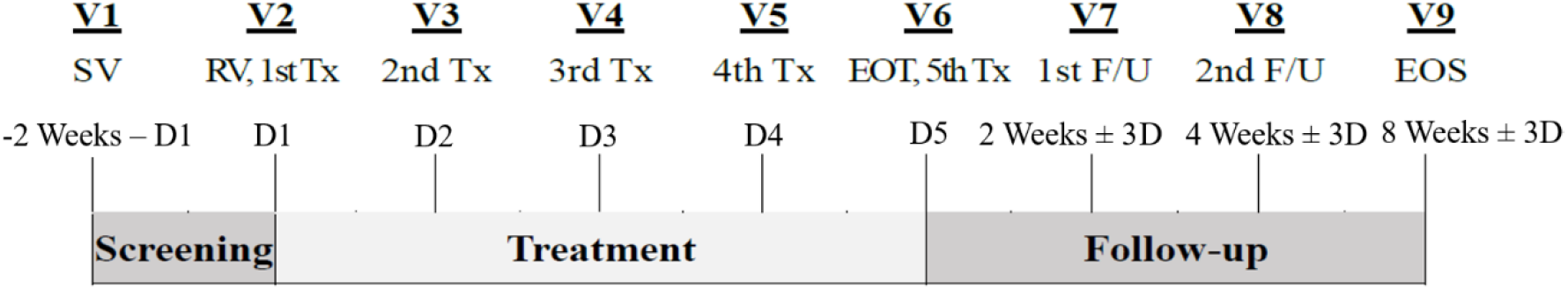
Study design. D, day; EOS, end-of-study; EOT, end-of-treatment; F/U, ; RV, ; SV, ; Tx, ; V, visit.

During the treatment period (V2–V6), the participants were monitored for safety and efficacy. The ischemic and leakage indexes were first measured at baseline (V1) and subsequently at 2, 4, and 8 weeks after administration (V7, V8, and V9, respectively).

### 2.6. Evaluation criteria

The primary endpoint was the mean percent change in ischemic index at each assessment point compared to baseline.

Secondary endpoints were: (i) fraction of participants who progressed to PDR (ETDRS ≥61) at each evaluation time point; (ii) DME-related changes at each assessment point compared to baseline (no DME, non-clinically significant macular edema); (iii) changes in CSF thickness and retinal volume assessed by spectral-domain optical coherence tomography at each assessment point; (iv) best-corrected ETDRS visual acuity measured at each assessment time point; (v) proportions of eyes that changed ≥1 stage and ≥2 stages (both improved and worsened) from baseline on the Diabetic Retinopathy Severity Scale (DRSS) at each time of assessment; (vi) fraction of eyes that changed ≥3 steps (both improvement and deterioration) from baseline on the DRSS at each time of evaluation; (vii) vascular leakage evaluated by UWFA at each time point; and (viii) changes in severity and frequency of peripheral lesions on UWFA and fundus examination measured at each assessment time point compared to baseline.

Safety endpoints were: (i) incidence and characteristics of adverse events after study drug administration (e.g., event pattern, severity, outcome); (ii) incidence and characteristics of prespecified adverse events (ocular adverse events); and (iii) physical examination, vital signs, electrocardiogram, and clinical laboratory examination findings related to the administration of the investigational drug.

### 2.7. Statistical analysis

Safety assessment group: This refers to the group of patients who received at least one dose of the investigational drug after being assigned to this trial and have undergone safety-related follow-up.

Full assessment (FA) group: All trial participants were eligible, but it refers to the group of patients who received at least one dose of the trial drug and whose drug efficacy can be assessed. The statistical analysis of the primary endpoint of this study was performed as a two-tailed test with a significance level (α) of 0.1; all other analyses were based on two-tailed tests with an α of 0.05.

Data were determined for the number of trial participants, incidence rates, and numbers of adverse events related to the investigational drug, as well as their severity, measures taken, and outcomes. The numbers of trial participants by system organ class (SOC) and preferred term (PT), incidence rates, and numbers of occurrences were summarized. Descriptive statistics of vital signs and quantitative laboratory tests provided the amount of change before and after investigational drug administration. Changes within the administration group were tested for differences compared to baseline using the paired t-test or Wilcoxon signed-rank test. Changes in normal/abnormal in physical, laboratory, and electrocardiogram examination before and after drug administration were summarized in a contingency table, and the significance of changes within the drug administration group was statistically tested using McNemar’s or Bowker’s test.

To further analyze EG-Mirotin effects, the V1 cutoff indicating a significant decrease in the ischemic index was determined using the ‘maxstat’ R package, and two groups were defined based on this cutoff. The repeated-measures ANOVA test was used to identify changes in ischemic and leakage indexes over time. To assess the difference in average ischemic index between these two groups, the Wilcoxon test, repeated-measures ANOVA, and Bonferroni post hoc analysis were used. R version 4.0.4 (R Project for Statistical Computing, Vienna, Austria), SPSS version 28 (IBM, New York, USA), and SAS version 9.4 (SAS Institute, North Carolina, USA) were used for statistical analyses and graphs generation.

## 3. Results

### 3.1. Study population

A total of 10 subjects were selected under the condition they were >19 years old, had type 1 or type 2 diabetes, and moderate-to-severe NPDR. Furthermore, media clarity should allow UWFA. Patients were excluded if they met at least one criterion that could affect the data interpretation. The average interval between visits in the treatment period was 23.22±3.32 days between baseline (V1) and V2, 12.12±3.36 days between V2 and V3, and 31.89±5.52 days between V3 and V4.

### 3.2. Safety evaluation

Adverse reactions: Four treatment-emergent adverse events (TEAEs) were observed in 2 out of 10 participants (20.00%) who received the investigational drug. These TEAEs were not related to the investigational drug. No adverse events leading to death were reported. One patient (10.00%) developed other serious adverse events but serious adverse reactions with a causal relationship to the investigational drug were not reported. None of the TEAEs resulted in drug discontinuation. One participant (10.00%, 2 cases) reported an adverse event defined in the study protocol (ocular adverse events: SOC eye disorders or ophthalmic procedures between surgical and medical procedures). All adverse reactions scheduled for collection did not correspond to TEAEs related to the investigational drug. As no adverse reactions were observed, the safety of the investigational drug was considered very good.

### 3.3. Drug efficacy

The primary efficacy outcome, the overall average percent change in ischemic index at each evaluation point compared to baseline, was statistically significant (Greenhouse-Geisser F=9.456, p=0.004 for the main effect of time), and a larger change was observed when the baseline value was high (Greenhouse-Geisser F=10.946, p=0.002 for time × group interaction). The baseline (V1) ischemic index for the 20 enrolled eyes was 3.82±4.78%, and the corresponding indexes at V7, V8, and V9 after study drug administration were 3.50±4.66%, 3.59±4.24%, and 3.11±3.85%, respectively. The changes from baseline were -0.28±0.46%, - 0.52±0.76%, and -0.71±1.02% (V7: p=0.0025, V8: p=0.0095, and V9: p=0.0005, respectively). When the baseline ischemic index was >1.59%, a statistically significant improvement over time was observed (>1.59 group: Wilk’s lambda F=9.142, p=0.002 for the simple main effect of time). By contrast, no improvement or deterioration was observed for patients with a low ischemic index value at baseline.

### 3.4. Secondary efficacy outcomes

Progression to PDR (ETDRS severity ≥61) was not observed. Changes in DME degree (without age-related macular degeneration (AMD), non-clinically significant macular edema): (i) most changes were not observed from baseline; (ii) 2 of 6 eyes (33.33%) assessed as senile macular degenearion (SMD) at baseline improved to grade ‘no SMD’ after treatment (V8 and V9); (iii) none of the eyes had a worse edema degree; and (iv) no significant changes in CSF thickness and retinal volume were observed between time points before and after drug administration; and (v) no significant changes in best-corrected ETDRS visual acuity were observed compared to baseline.

Changes in DRSS scores from baseline: (i) no eye improved 1 or 2 steps; (ii) one eye was categorized as deterioration of 1 or 2 steps; and (iii) no eye had ≥3 steps (improvement or deterioration).

Change in vascular leakage rate from baseline: The average leakage index decreased significantly over time (Greenhouse-Geisser F=6.442, p=0.008 for the overall main effect of time). After test drug administration, changes at V7, V8, and V9 were significantly different with -0.69±1.17%, -0.85±1.53%, and -2.09±2.32% (V7: p=0.0233, V8: p=0.0305, and V9: p=0.0001, respectively).

Changes in the severity and frequency of peripheral lesions: (i) Retinal hemorrhage/retinal microvascularization: ‘no change’ in severity in 18 eyes (90.00%), ‘Best’ in 2 eyes (10.00%), and no changes in the distribution; (ii) venous beading: ‘no change’ in severity compared to baseline for all V9 visits (100.00%), one eye each with dilated venous beading with intraretinal and superonasal microvascular abnormalities; and (iii) severity: ‘unchanged’ in 18 eyes (90.00%), ‘Best’ in 2 eyes (10.00%), and no changes in the distribution

### 3.5. Ischemic index

The UWFA images of 17 eyes from 9 patients were assessed before (V1, baseline) and 2, 4, and 8 weeks after (V7, V8, and V9, respectively) drug administration. There was an overall significant decrease in ischemic indexes across visits (Greenhouse-Geisser F=9.456, p=0.004 for the main effect of time). The average ischemic index decreased over time, except for the interval between V7 and V8 (Fig. 3). EG-Mirotin administration significantly decreased the average ischemic index values from baseline (3.82±4.78) to V9 (3.11±3.85; p=0.0005).

**Fig. 3.**
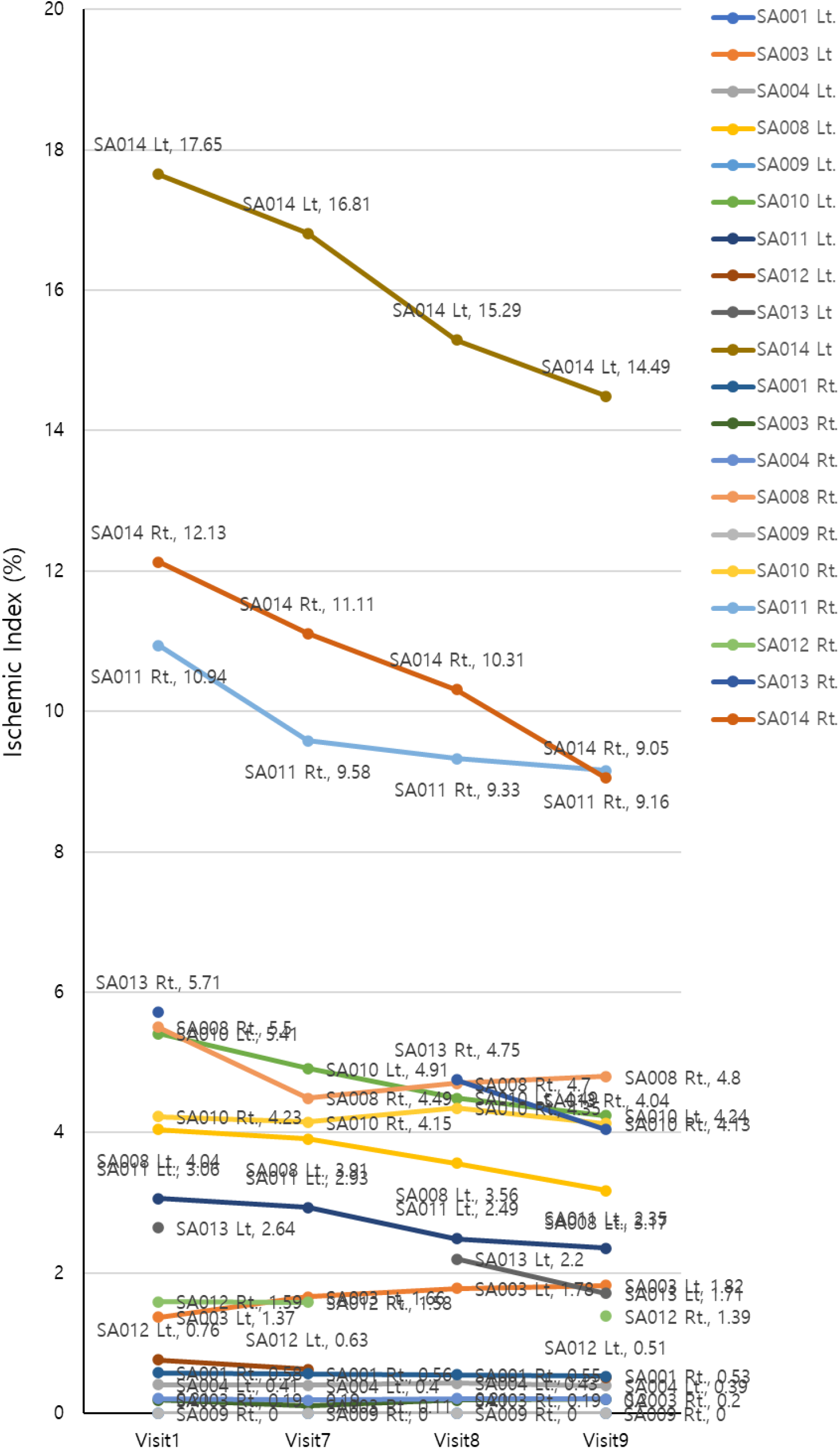
Changes in ischemic indexes of the participants during the study period.

The ischemic percent difference (IPD) was defined as the differences in ischemic indexes between V1 and V7 (IPD1, Fig. 4A), V1 and V8 (IPD2, Fig. 4B), and V1 and V9 (IPD3 Fig. 4C). Patients were divided into two groups based on an ischemic index cutoff of 1.59% at baseline. All eyes with an ischemic index >1.59 decreased in size (Figs. 4 & 5). The two groups were significantly different regarding IPD1 (p<0.001), IPD2 (p=0.004), and IPD3 (p<0.001, Wilcoxon test; Fig. 4).

**Fig. 4.**
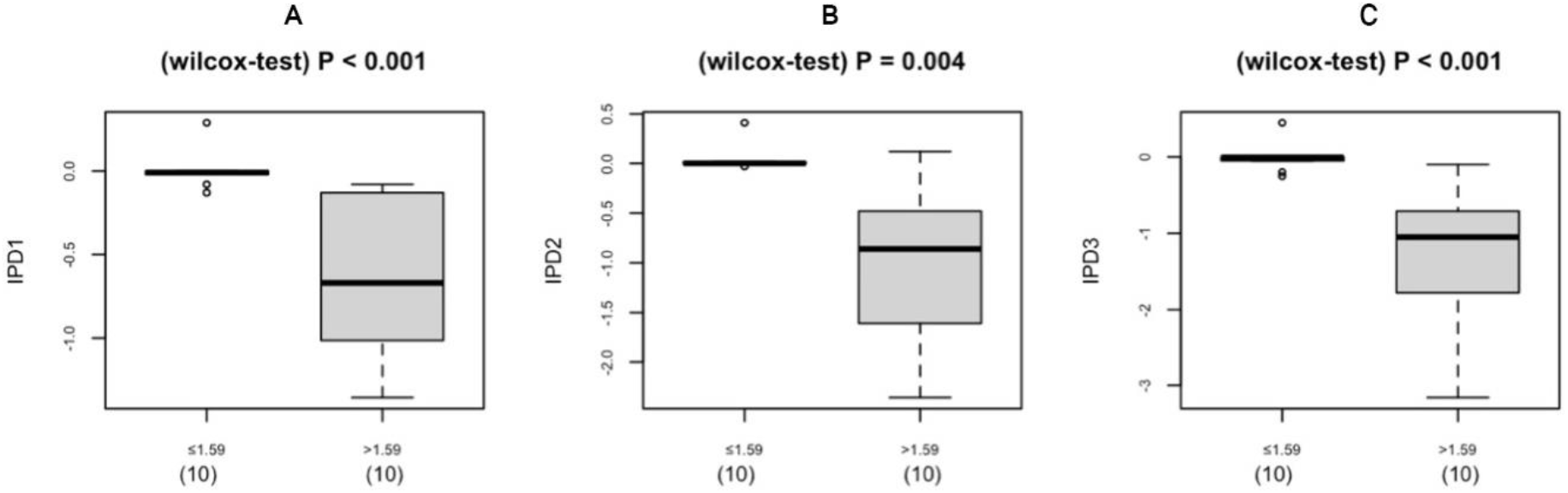
Boxplot of the IPD distribution for eyes below (left) and above (right) the cutoff of 1.59% at baseline (V1). A: IPD1 (V1 vs. V7); B: IPD2 (V1 vs. V8); C: IPD3 (V1 vs. V9). IPD, ischemic percent difference; V, visit.

**Fig. 5.**
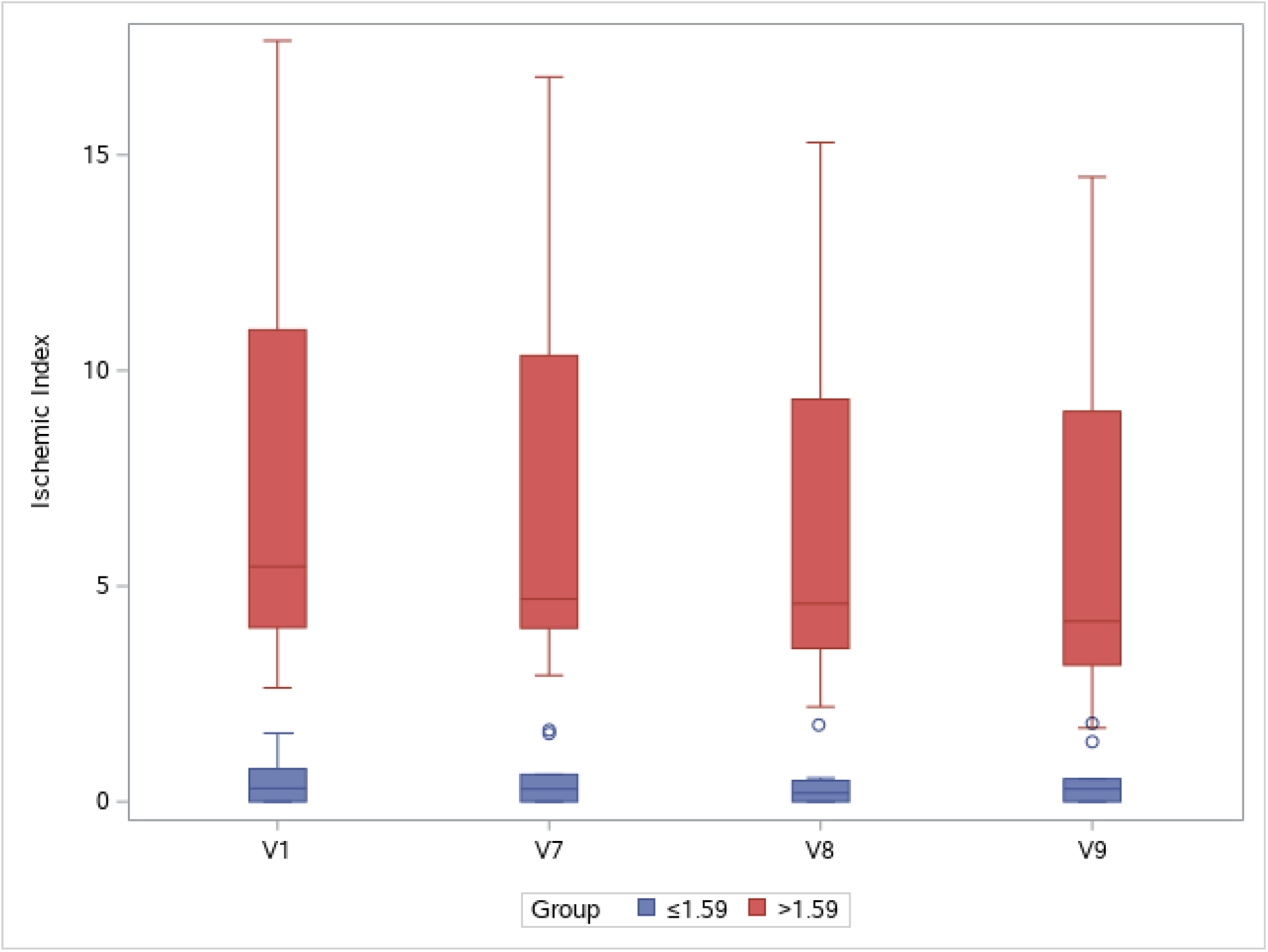
Changes in ischemic indexes throughout the study period. Groups are separated by a cutoff value of 1.59% at baseline (V1). V, visit.

**Fig. 6.**
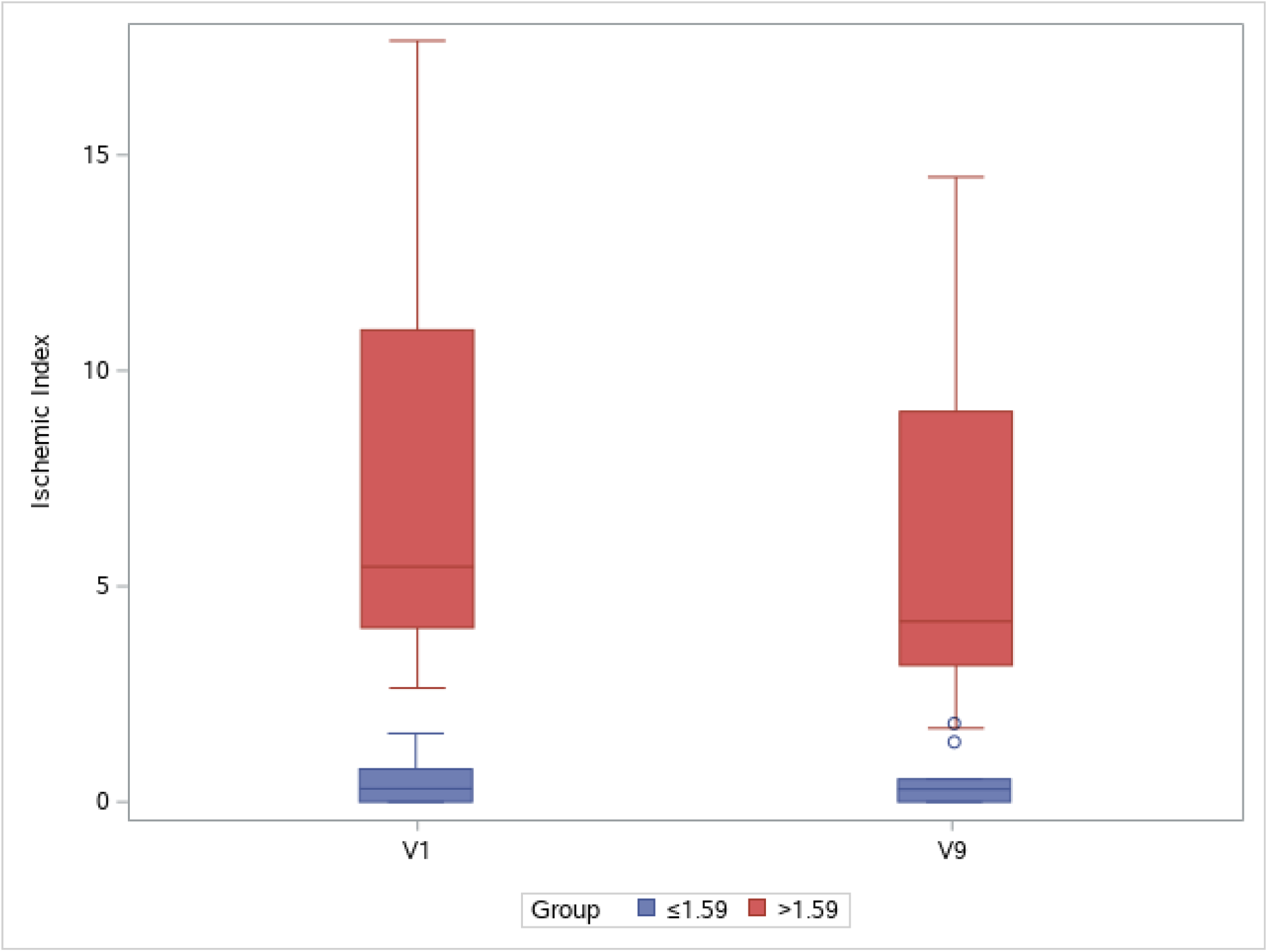
Changes in ischemic index between V1 and V9. Groups are separated by a cutoff value of 1.59% at baseline (V1). V, visit.

To observe changes in ischemic indexes within each group, the repeated-measures ANOVA test was used. The ischemic index significantly decreased over time (Greenhouse-Geisser F=9.456 for the main effect of time), and a significant interaction effect was observed (Greenhouse-Geisser F=10.946 for time × group interaction). A significant change in ischemic index was found in the >1.59 group (Wilk’s lambda F=9.142, p=0.002 for the simple main effect of time), whereas no change was observed in the ≤1.59 group (Wilk’s lambda F=0.038, p=0.990 for the simple main effect of time).

In the >1.59 group, the ischemic index was significantly different compared to the baseline value at V7, V8, and V9 (p=0.001, p<0.001, and p=0.001, respectively; Bonferroni post hoc test). For the ≤1.59 group, the ischemic index did not significantly differ among visits. In the >1.59 group, EG-Mirotin administration decreased the average ischemic index from 7.13±4.86 at baseline to 5.71±3.97 at V9. Thus, for patients with an ischemic index above the 1.59% cutoff, EG-Mirotin administration significantly decreased the ischemic index value.

### 3.6. Leakage index

The UWFA images of 20 eyes from 10 patients were assessed before (V1, baseline) and 2, 4, and 8 weeks after (V7, V8, and V9, respectively) drug administration. There was an overall significant decrease in leakage indexes across visits (Greenhouse-Geisser F=6.442, p=0.008 for the main effect of time). The average leakage index decreased over time, except for the interval between V7 and V8. Although the mean value increased between V7 and V8, this change was not statistically significant (p=0.14). EG-Mirotin administration decreased the average leakage index from baseline (13.60±11.21) to V9 (11.51±9.80, p=0.0001; Fig. 7).

**Fig. 7.**
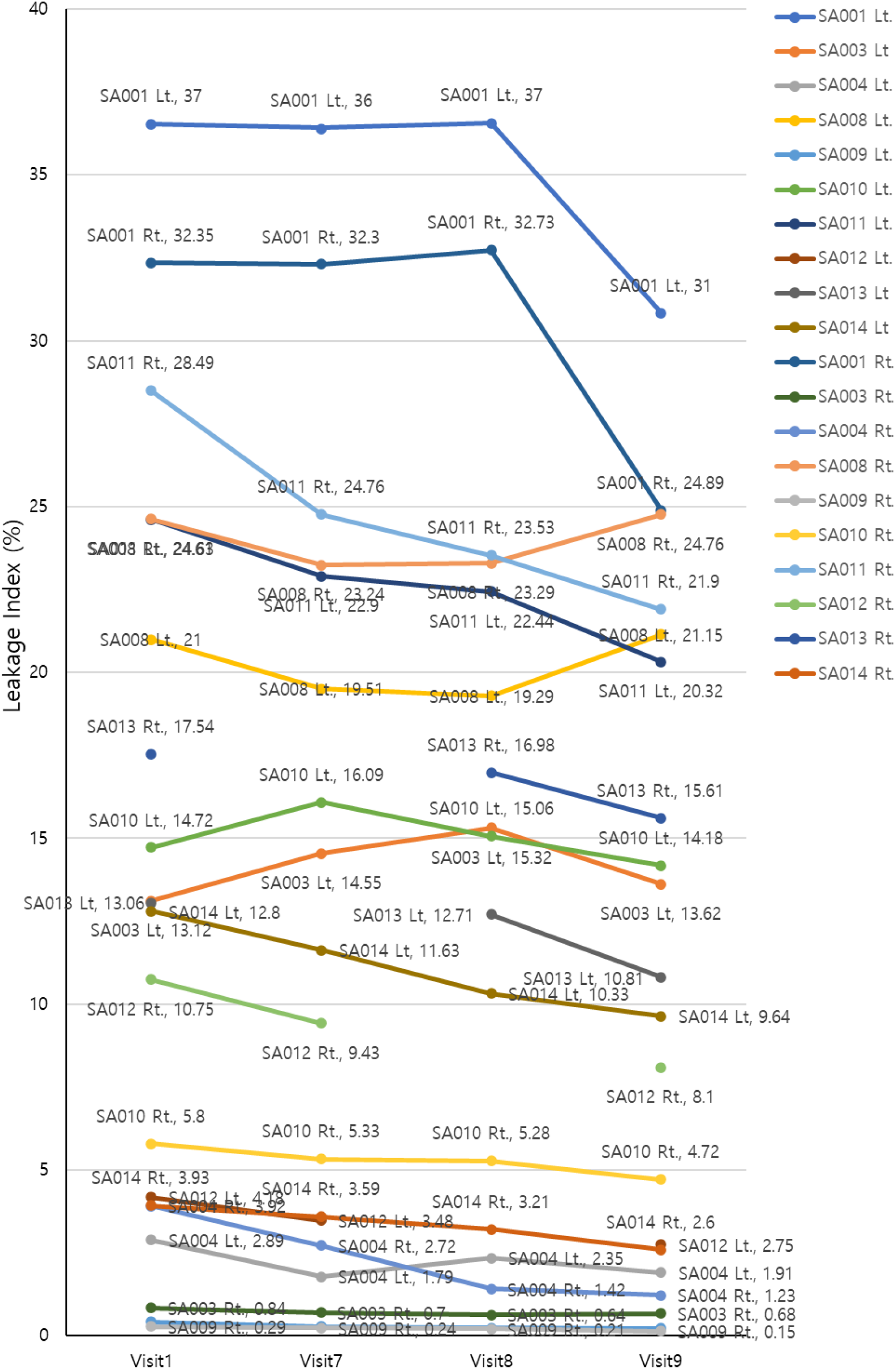
Changes in leakage indexes of the participants throughout the study period.

To identify whether ischemic and leakage indexes were correlated at each time point, Spearman’s rank correlation test was conducted, as these indexes were not normally distributed (Shapiro-Wilk w=0.7868, p<0.0001 for ischemic index and w=0.9167, p<0.0001 for leakage index). The two indexes were moderately correlated (Spearman’s ρ=0.3779, p=0.0007; Fig. 8).

**Fig. 8.**
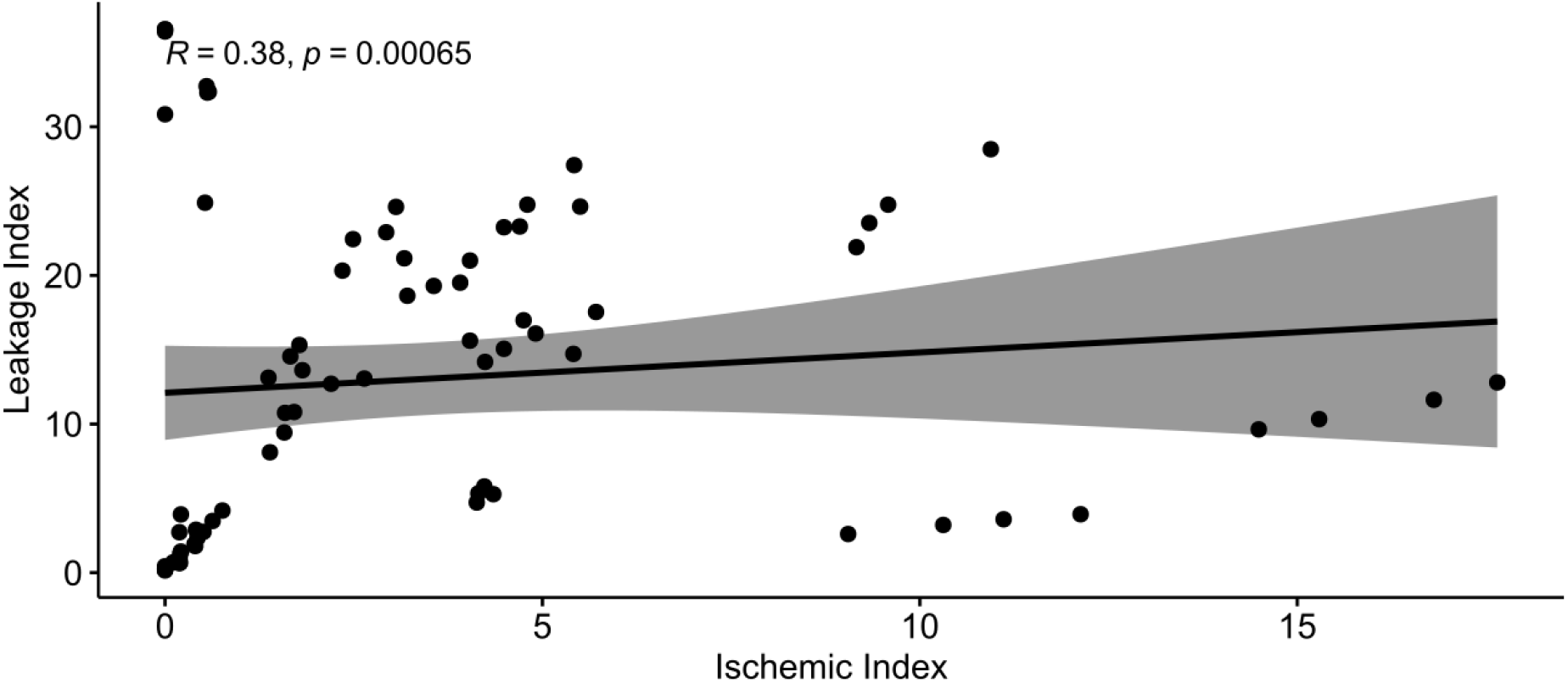
Spearman’s correlation between ischemic and leakage indexes.

## 4. Discussion

Diabetes mellitus is perhaps the most persistent disease in human history. Despite the tremendous achievements of diabetes research, this disease still negatively affects patient well-being with numerous complications in varying loci and severity grades. DR, a common but severe ocular complication, is a major cause of blindness and, thus, greatly diminishes the quality of life over a long time. Currently, there is no approved drug for early-stage DR, i.e., NPDR, and patients are without treatment until the disease has progressed to PDR. We believe that improving the quality of life of patients with NPDR by slowing or halting the progression of visual impairment with an early-stage intervention is a challenge in modern medicine.

Recent DR treatment trends have two main therapeutic goals: (i) prevention of progression from NPDR to PDR, which has a high risk of visual loss due to vitreous hemorrhage or tractional retinal detachment and (ii) prevention of loss of central vision due to DME. To achieve these goals, it is necessary to prevent capillary leakage or suppress neovascularization caused by increased VEGF levels in the eye due to retinal ischemia. PRP, which has been used for a long time, effectively lowers VEGF levels. However, PRP irreversibly destroys the retina to prevent NPDR reoccurrence afterward [16]. The recently popular method, intraocular injection of anti-VEGF antibodies, is a potent therapy that can achieve these goals more quickly. However, this approach aims at short-term effects in an irreversible disease stage where blood vessels cannot return to a normal state and the retinal condition continues to gradually deteriorate. If it were possible to reduce retinal capillary damage and related retinal ischemia, which are the causes of capillary leakage and neovascularization, the risk of blindness could be reduced by preventing the progression of DME and PDR.

EG-Mirotin was developed to help maintain normal blood vessels in patients with DR before irreversible capillary damage occurs. Diabetes is a disease in which high levels of circulating blood sugar damage blood vessels. In the human body, the kidneys and retina are the organs with the highest number of capillaries and the highest blood flow per unit area. Therefore, these organs are mainly affected by diabetes, and retinal damage ultimately increases the risk of blindness. In diabetes, abnormally high blood sugar levels can damage endothelial cells, which in capillaries only have one cell layer. For about 5 years of diabetes, cells surrounding the capillaries restore this endothelial cell layer and ensure normal capillary function. Afterward, even these surrounding cells are damaged, and capillary microaneurysms form, causing vessel occlusions and hemorrhages. Perivascular cells vanish, capillaries are irreversibly damaged, the retina cannot maintain its normal morphology, and DR eventually progresses regardless of treatment. With the expansion of retinal ischemic areas, VEGF levels increase to stimulate the development of new retinal capillaries counteracting ischemia. If it were possible to maintain or revive the surrounding cells at the stage of endothelial cell damage, retinal capillaries could be rescued, and ischemia could be reduced.

EG-Mirotin was developed to prevent retinal damage and blindness by restoring capillaries before their damage progresses to an irreversible stage. Existing anti-VEGF antibodies for intraocular injection reduce damage to new blood vessels in the irreversible stage. Although aflibercept (Eylea®), an anti-VEGF treatment, is also approved for NPDR in the USA, the need for frequent ocular injections burdens patients and physicians, thus precluding accessibility at the NPDR stage. EG-Mirotin was developed to prevent capillary damage as early as possible in the NPDR stage so that the disease does not progress to an irreversible stage with the additional advantage of subcutaneous administration. The findings of this study confirmed that EG-Mirotin administration restored the capillaries where retinal ischemia occurred, thereby reducing the ischemic area in the retina. By blocking irreversible capillary damage via recruitment of surrounding cells and actively initiating treatment at the NPDR stage, progression to the proliferative stage can be largely prevented. As EG-Mirotin is subcutaneously administered instead of into or around the eye, the patient’s fear of ocular injection is reduced. Since the mechanism of this drug constitutes a novel approach, research data is continually accumulating. In the future, more research will be conducted on EG-Mirotin to develop better drugs for patients with NPDR.

In the present study, progression to proliferative DR did not occur in any of the participants. Moreover, both ischemic and leakage index values determined in UWFA images of lesions related to fundus microvascularization were significantly reduced compared to baseline values. Of note, a greater degree of change was observed in patients who had a larger ischemic index value at baseline. The participants were further categorized into two groups based on the cutoff of 1.59% in the baseline ischemic index. There was a significant interaction between group assignment and therapeutic effect over time, of which statistical significance was only observed in the >1.59 group in the post hoc comparison. It is expected that the ≤1.59 group showed no difference in the ischemic index over time because there was little room for change as the initial ischemic value was ≤1.59% with a few patients having an ischemic index of zero. The lack of index change can also be interpreted as the absence of disease deterioration. As retinal ischemia is a major factor in the pathogenesis of PDR [17], the prevention of ischemia may protect patients from progression to PDR. A larger placebo-controlled follow-up study with longer observation duration will be required to confirm the protective effects of EG-Mirotin.

EGT022 is a protein containing an arginine-glycine-aspartic acid sequence [18], and our in-house experiments provided novel data: (i) EGT022 restores retinal blood vessels in the OIR mouse model (data not shown) and (ii) EGT022-induced maturation of functional retinal blood vessels is closely associated with pericyte recruitment and blood vessel integrity [8]. It can be assumed that the significant reduction in ischemia and leakage can be attributed to the active ingredient of EG-Mirotin and its molecular mode of action of pericyte recruitment [19], thereby stabilizing blood vessels.

We found no serious unwanted effects or safety concerns following EG-Mirotin administration. None of the participants reported TEAEs that resulted in premature drug discontinuation. In preclinical studies, EG-Mirotin had no acute toxicity or genotoxicity. EG-Mirotin alone did not induce angiogenesis and had no growth-promoting effects on malignancies. The efficacy of EG-Mirotin in retinal edema has been demonstrated in the Miles assay as a supplemental assay to assess the vascular permeability of microvessels. EG-Mirotin appeared to be effective in ameliorating retinopathy-related abnormalities for up to 3 weeks as demonstrated by optical coherence tomography, fundoscopy, angiography, and histopathology in a streptozotocin-induced rat model of DR [8].

Our study has several limitations. Based on our preclinical experience, we chose to use 3 mg of EG-Mirotin, which may not be a sufficient dose to cause substantial changes in retinal blood vessels. Theoretically, the dose could be increased to 10 mg per administration. However, we need to collect more safety data to justify a dose increase. In the present study, the participants visited the clinic during the treatment period (i.e., a total of 5 days: V2–V6). A daily clinic visit is a nuisance for many patients. Therefore, our future goal is to prepare EG-Mirotin for self-injection, as established for insulin administration, so patients do not have to visit daily the clinic for their drug treatment.

## 5. Conclusion

We developed EG-Mirotin (EGT022), a novel, first-in-class, subcutaneously delivered peptide drug for NPDR, for which currently no selectively designed drug exists. The findings of this clinical trial suggest the safety and efficacy of EG-Mirotin, as no serious adverse events with a causal relationship to EG-Mirotin were reported during the study period, and an overall improvement of ischemic and leakage indexes with no progression to PDR was observed. To further verify the therapeutic effects of EG-Mirotin on DR, larger clinical trials focusing on patients with high baseline ischemic indexes are required.

In conclusion, EG-Mirotin has provided new hope for the treatment of NPDR which, so far, has had few treatment options. In addition to being a selectively designed first-in-class drug for NPDR, EG-Mirotin is also subcutaneously administered, reducing the burden and fear associated with intraocular drug injection, which is currently the only available treatment for PDR, the late, irreversible stage of DR. Moreover, EG-Mirotin was found to reverse retinal ischemia and capillary leakage caused by diabetes. We hope that EG-Mirotin will preserve the well-being of patients with DR at the reversible stage by greatly reducing the risk of loss of vision with the additional benefit of simple drug administration.

## Data Availability

All data produced in the present study are available upon reasonable request to the authors

## Author contributions

Study conception and design: SY, SJL

Data acquisition: SY, SJL

Data analysis and interpretation: SY, DHY, JL, HCH, SJL

Review and final approval of the manuscript: SY, DHY, JL, HCH, SJL

## Clinical Trial Registration

Clinical Research Information System (CRIS): KCT0005124

The above KCT0005124 has been listed on the International Clinical Trials Registry Platform of the World Health Organization

https://trialsearch.who.int/Trial2.aspx?TrialID=KCT0005124

## Clinical Trial (IRB Approval Number)

SCHUH2020-03-004

